# Validation of Pressure-Strain Loops for Non-Invasive Assessment of Ventriculo-Arterial Coupling

**DOI:** 10.64898/2026.03.08.26347879

**Authors:** Lígia Mendes, João Colaço, José Ferreira Santos, João Pereira, Ana Teresa Timóteo

## Abstract

**Background and Objectives:** Left ventricular pressure–strain loop (LV-PSL) analysis provides noninvasive myocardial work indices that may reflect ventricular–arterial (VA) coupling, but their behavior under controlled physiologic stressors is incompletely defined. We aimed to characterize directional changes in LV-PSL, derived indices during standardized interventions predominantly affecting preload, afterload, or contractility in healthy adults.

**Methods:** In this prospective, within-subject repeated-measures study, 61 healthy volunteers underwent interventions designed to elicit domain-specific hemodynamic perturbations. Group 1 (n=31) performed isotonic exercise (contractility-dominant). Group 2 (n=30) performed isometric handgrip (afterload-increasing) and passive leg raising (PLR; preload augmentation with concurrent afterload change). LV-PSL indices were assessed at baseline and during each intervention. Six co-primary endpoints were prespecified: Global Work Index (GWI), peak systolic strain, strain range, systolic strain rate (SSR), arterial elastance (Ea), and end-systolic pressure (ESP). Within-subject changes were analyzed using paired tests with multiplicity adjustment and determined effect sizes. Reproducibility was evaluated using intraclass correlation coefficients (ICC).

**Results:** LV-PSL responses were directionally consistent with established pressure–volume physiology. Isotonic exercise produced large increases in contractility-sensitive indices, including GWI (dz=1.03), peak systolic strain (dz=0.88), strain range (dz=1.10), SSR (dz=1.29), and ESP (r=1.26), all adjusted p<0.001, while Ea remained unchanged. In contrast, isometric handgrip and PLR elicited afterload-dominant responses, with significant increases in ESP (dz=1.11 and 1.21, respectively; adjusted p<0.001) and Ea (dz=0.79 and 0.77; adjusted p≤0.001), without significant changes in GWI or strain-derived indices after adjustment. Intraobserver reproducibility was good-to-excellent (ICC 0.86–0.90), and interobserver reproducibility was moderate-to-good (ICC 0.72–0.87).

**Conclusions:** In healthy adults, LV-PSL indices demonstrate good reproducibility and appropriate sensitivity to hemodynamic perturbations. Exercise elicited contractility-dominant responses, whereas handgrip and PLR induced afterload-dominant changes. These physiologically coherent response patterns support LV-PSL as a practical non-invasive surrogate for invasive pressure–volume assessment.

Central Illustration - Validation of Non-Invasive Pressure-Strain Loops for Assessing Ventriculo-Arterial Coupling
The study evaluated left ventricle pressure-strain loop (LV-PSL) derived indices during three hemodynamic interventions in healthy volunteers: Group 1 - exercise (contractility-dominant), Group 2 - isometric handgrip (afterload-dominant), and passive leg raising (preload/afterload modulation). Center heatmap shows effect sizes (Cohen’s dz or rank-biserial r) for six co-primary PSL endpoints. Color intensity indicates effect magnitude (red = positive, blue = negative); asterisks denote significance after Holm-Bonferroni correction (**p≤0.001).
Exercise produced robust responses in 5/6 parameters, while handgrip and passive leg raising showed intervention-specific patterns, particularly for afterload indices.
PSL methodology demonstrates high reproducibility and physiological sensitivity for non-invasive ventriculo-arterial coupling assessment.
**Abbreviations:** LV-PSL, Left ventricle pressure-strain loop; PLR, passive leg raising; VA, ventriculo-arterial

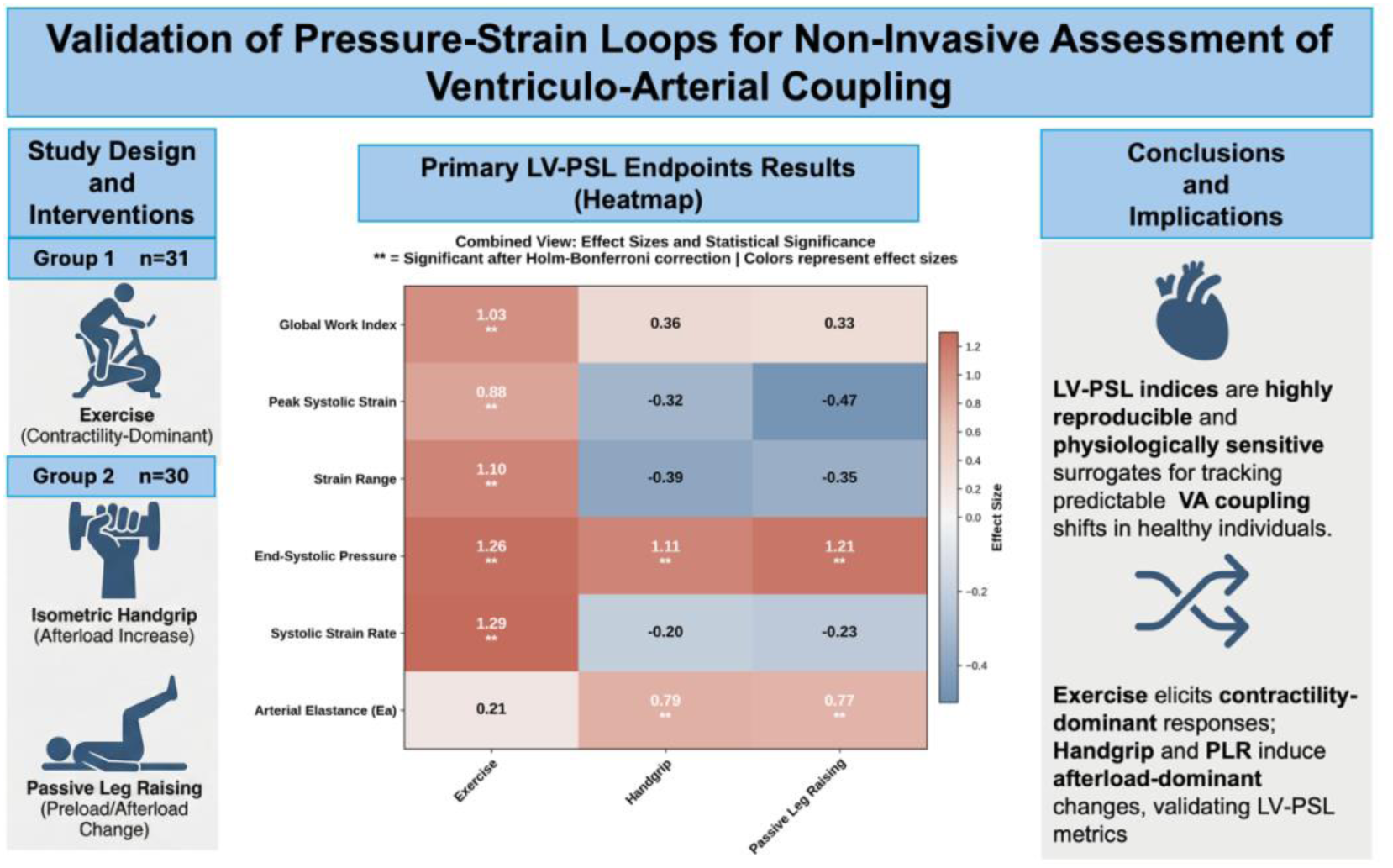

## Introduction

The interaction between the left ventricle (LV) and the arterial system - termed ventriculo-arterial (VA) coupling - is a fundamental determinant of cardiovascular performance. Based in Otto Frank’s seminal pressure–volume (PV) framework, VA coupling describes how the heart’s contractile properties adapt to the load imposed by the arterial tree to preserve efficiency and optimize stroke work (1–3). Quantifying this interaction provides an integrated measure of cardiac energetics and mechanical performance, both under physiological conditions and in diseases (4–6).

The invasive PV loop remains the reference standard for assessing LV mechanics and VA coupling, enabling derivation of key indices such as end-systolic elastance ventricular (Ees) and effective arterial elastance (Ea). However, this technique requires conductance catheterization, meticulous calibration, and specialized expertise, rendering it impractical for routine use or serial assessments (7,8).

To overcome these limitations, Russell et al. introduced the left ventricular pressure–strain loop (LV-PSL) analysis, which combines an estimated LV pressure curve (scaled from brachial cuff pressure and echocardiographic valve timing), with myocardial strain obtained by speckle-tracking echocardiography (9). The enclosed loop area represents the Global Work Index (GWI), a measure of myocardial work expressed in mmHg*%. This non-invasive index correlates strongly with myocardial oxygen consumption and metabolic demand, providing a physiologically meaningful surrogate of regional cardiac work. Further validation studies have demonstrated that PSL-derived indices, such as Global Constructive Work and GWI, correspond closely with invasively measured parameters, even when absolute pressure estimates vary modestly (10).

Despite its promise, the application of LV-PSL as a non-invasive tool for dynamic VA coupling assessment remains incompletely validated across physiological stressors. The present proof-of-concept study aims to validate non-invasive LV-PSL indices as surrogates for pressure-volume physiology by quantifying their response to standardized preload, afterload, and contractility stressors, hypothesizing that these metrics will reflect predictable hemodynamic shifts (6,7).

## Methods

### Study design and participants

This was a prospective, within-subject, repeated-measures study in healthy adult volunteers. Exclusion criteria included hypertension, diabetes mellitus, dyslipidemia, current smoking, pregnancy, significant systemic illness, use of cardioactive medications (agents affecting heart rate, contractility, or vascular tone), and baseline echocardiographic abnormalities suggestive of structural heart disease.

Participants were allocated to one of two standardized intervention protocols designed to elicit predominantly domain-specific hemodynamic perturbations. Group 1 underwent semi-supine isotonic exercise (contractility-dominant). Group 2 performed isometric handgrip (predominantly afterload-increasing) followed by passive leg raising (PLR; preload augmentation with concurrent afterload change). In Group 2, a 5-minute recovery period was maintained between maneuvers. PLR was performed with the legs supported on a platform in a tabletop position with flexed knees.

All interventions were conducted in a controlled laboratory setting with contemporaneous brachial blood pressure acquisition. Echocardiographic imaging was obtained at physiologic steady state during handgrip, within the established post-PLR response window, and at a prespecified exercise workload/heart-rate target (>25 Watts and approximately 100 beats/min) during semi-supine exercise.

As prespecified, analyses were restricted to participants with paired baseline and intervention acquisitions of adequate-to-optimal image quality for deformation (speckle-tracking) analysis. Enrollment was concluded once at least 30 valid paired acquisitions were obtained for each protocol.

### Echocardiographic acquisition and PSL construction

Transthoracic echocardiography was obtained in apical two-, three-, and four-chamber views and analyzed offline in EchoPAC v204 (GE Healthcare®). Global longitudinal strain (GLS) was derived using speckle-tracking according to the methodology of the contemporary consensus (11). A left-ventricular pressure waveform was estimated by scaling a reference curve to brachial systolic/diastolic pressures and aligning it to mitral and aortic valve timing. LV-PSL-derived indices included myocardial work parameters (global work index (GWI), global constructive work (GCW), global wasted work (GWW), and global work efficiency (GWE)) and strain-based measures (peak systolic strain, strain range, systolic strain rate (SSR), end-diastolic strain, and early diastolic strain rate), as well as filling time, diastolic time, and ratios (11–14).

The effective arterial elastance surrogate (Ea, mmHg/%) was derived analogously to pressure-volume loop methodology, where Ea = end-systolic pressure / stroke volume. In the pressure-strain domain, Ea was calculated as the ratio of estimated end-systolic pressure (ESPest) to global longitudinal Δε (strain range; peak systolic strain − end-diastolic strain): Ea = ESPest / Δε. End-systolic pressure was estimated as 0.9 × brachial systolic blood pressure. This parameter represents the slope of the line from the origin to the end-systolic point in pressure–strain space and serves as a non-invasive index of afterload.

### Endpoints

Six co-primary endpoints were pre-specified to capture distinct hemodynamic properties: Ea and end-systolic pressure (ESP), afterload; systolic strain rate (SSR) and peak systolic strain, contractility; and strain range and GWI, work/energetics. To assess the robustness of findings, sensitivity analyses were performed on all primary endpoints across interventions.

Secondary endpoints comprised qualitative loop-morphology assessment, directional and shape changes of LV-PSL pre- versus post-intervention, obtained by superimposing paired loops, and were interpreted against established pressure–volume behavior. Additional secondary measures included myocardial-work components (constructive/wasted work and global work efficiency) and ancillary timing indices.

### Expected physiological directions (*a priori*)

Directional hypotheses for each endpoint under each intervention were defined *a priori* based on invasive PV loop data (**Supplementary Table S1**). Handgrip, as an afterload-augmenting maneuver, was expected to increase estimated end-systolic pressure and the Ea surrogate, with minimal or modest reductions in peak systolic strain. Exercise was hypothesized to increase systolic strain rate, augment peak systolic strain, and raise myocardial work indices, consistent with enhanced inotropy. PLR was anticipated to elicit a mixed hemodynamic profile rather than a purely preload-driven shift, with a preload-mediated increase in loop dimensions (GWI and loop width) accompanied by an afterload rise (Ea and end-systolic pressure) attributable to increased peripheral resistance in the flexed-leg position (15–17).

### Data handling and reproducibility

EchoPAC exports (xml) were parsed using Python 3 to Pandas 2 data frames and exported to an Excel file. Custom Python utilities also generated superimposed LV-PSL to quantify loop-shape secondary endpoints. Strain values are expressed in absolute numbers.

### Statistical analysis

The Statistical Analysis Plan required at least 30 paired cases per intervention group; all power and precision calculations are detailed therein. Within-subject change scores (Δ = intervention − baseline) were tested per endpoint and intervention using the Shapiro–Wilk test for normality, followed by paired t-tests (normal distribution) or exact Wilcoxon signed-rank tests (non-normal distribution). Effect sizes were reported as Cohen’s dz (parametric) or rank-biserial r (non-parametric) with 95% confidence intervals (CI), using standard thresholds: for Cohen’s dz, negligible (<0.2), small (0.2–0.5), medium (0.5–0.8), and large (>0.8); for rank-biserial r, negligible (<0.1), small (0.1–0.3), medium (0.3–0.5), and large (>0.5). Multiplicity was controlled within intervention families using Holm–Bonferroni across the six co-primary endpoints (family-wise α=0.05). Sensitivity analyses included outlier removal, Winsorization, permutation tests (1,999 sign-flips), and BCa (Bias-Corrected and accelerated) bootstrap CIs (1,999 resamples). Secondary endpoints were descriptive (p-values, effect sizes/CIs are only nominal). Reproducibility was assessed in a 15-participant subset using blinded duplicate reads with an ≥8-week washout to quantify intra- and inter-observer reliability (intraclass correlation coefficients with 95% CIs, ICC(3,1) for intra-observer and ICC(2,1) for inter-observer) (18) and agreement (Bland–Altman bias and limits with proportional-bias testing). Analyses were performed in R (RStudio version 2025.09.2+418).

### Ethics and registration

The study complied with the principles of the Declaration of Helsinki. All participants provided written informed consent before enrollment, and the protocol received institutional ethics approval (19). The project is publicly registered on the Open Science Framework (OSF): https://doi.org/10.17605/OSF.IO/3QJGW.

## Results

### Participant Characteristics

Recruitment continued until achieving 30 high-quality acquisitions per intervention: 56 volunteers were screened for exercise (31 analyzable) and 47 for Handgrip/PLR (30 analyzable). Importantly, the baseline demographic and hemodynamic characteristics of the participants included in the final analyses were not different from those of the total enrolled cohorts, indicating no significant selection bias (**Supplementary Table S2)**. Among included volunteers, women comprised 65% of the exercise cohort versus 47% of those undergoing bedside maneuvers. Mean age was similar between groups (42 ± 8 vs. 43 ± 8 years). Resting brachial blood pressure was comparable (systolic 122 ± 15 vs. 121 ± 20 mmHg; diastolic 72 ± 8 vs. 69 ± 9 mmHg), though BMI was higher in the exercise cohort (29.0 ± 4.7 vs. 25.1 ± 3.2 kg/m^2^).

The baseline hemodynamic and echocardiographic characteristics of the 61 participants included in the final analyses are presented in **Table 1**. Both the exercise and the maneuvers group demonstrated echocardiographic parameters consistent with healthy cardiovascular function. Global work efficiency exceeded 90% in both groups, and global work index values fell within the established normal range for healthy adults. End-diastolic strain, systolic strain rate, and early diastolic strain rate were all within expected physiological limits, confirming an appropriate healthy baseline before intervention.

**Table 1.** Baseline hemodynamic and echocardiographic characteristics. **Abbreviations:** Ea (arterial elastance); Filling time proportion (fraction [filling time/cycle length]); GCW (global constructive work); GWE (global work efficiency); GWI (global work index); GWW (global wasted work); IVRT (isovolumic relaxation time); IVRT proportion (fraction [IVRT time/cycle length]); Strain range (dimensionless [peak strain - ED strain]); Systolic time proportion (fraction [systole time/cycle length]).

| **Basal Characteristic** | **Group 1**<br>**Exercise**<br>**N = 31<sup>1</sup>** | **Group 2**<br>**Manoeuvres**<br>**N = 30<sup>1</sup>** |
| --- | --- | --- |
| **Estimated End Diastolic Pressure** (mmHg) | 8.84 ± 1.06 | 8.61 ± 0.67 |
| **End systolic pressure estimate** (mmHg) | 109 ± 13 | 106 ± 8 |
| **GWE** (%) | 90 ± 4 | 95 ± 2 |
| **GWI** (mmHg*%) | 1,413 ± 287 | 1,684 ± 276 |
| **GCW** (mmHg*%) | 1,780 ± 346 | 2,015 ± 312 |
| **GWW** (mmHg*%) | 179 ± 107 | 99 ± 43 |
| **End-diastolic strain** (%) | 0.043 ± 0.005 | 0.045 ± 0.006 |
| **Peak systolic strain** (%) | 14.93 ± 2.01 | 18.75 ± 2.67 |
| **Systolic strain rate** (s<sup>-1</sup>) | 0.72 ± 0.12 | 0.82 ± 0.10 |
| **Early-diastolic strain rate** (s<sup>-1</sup>) | 0.41 ± 0.20 | 0.68 ± 0.36 |
| **Strain Range** | 14.79 ± 1.98 | 18.35 ± 2.53 |
| **Systolic duration** (ms) | 279 ± 32 | 279 ± 28 |
| **IVRT** (ms) | 101 ± 4 | 100 ± 8 |
| **Filling Time** (ms) | 464 ± 141 | 572 ± 149 |
| **Total diastolic time** (ms) | 565 ± 142 | 672 ± 147 |
| **Systolic time proportion** | 0.34 ± 0.05 | 0.30 ± 0.04 |
| **IVRT proportion** | 0.123 ± 0.020 | 0.109 ± 0.022 |
| **Filling time proportion** | 0.54 ± 0.06 | 0.59 ± 0.05 |
| **Ea** (mmHg/%) | 7.49 ± 1.30 | 5.89 ± 0.98 |

<sup>1</sup> Mean ± SD

### Primary endpoints

Across the three interventions, within-subject changes followed the pre-specified physiological directions, and several reached significance after Holm–Bonferroni correction (see **central illustration**). Full change scores, effect sizes with 95% CI, and adjusted p-values are provided in **Table 2** and **Supplementary Table S3.**

**Table 2.** Primary endpoints within-subject change. 95% confidence intervals are unadjusted. Standardized effect size (Cohen’s dz or rank-biserial r). Statistical significance was determined using Holm-Bonferroni adjusted p-values to control the family-wise error rate across the six co-primary endpoints within each intervention. **Effect size**: Cohen’s |dz| < 0.2 (negligible), 0.2-0.5 (small), 0.5-0.8 (medium), >0.8 (large) and Matched-pairs rank-biserial |r| < 0.1 (negligible), 0.1-0.3 (small), 0.3-0.5 (medium), >0.5 (large) **Abbreviations:** PLR (passive leg raising)

| Intervention | Endpoint | Baseline (mean ± SD) | Post-maneuver (mean ± SD) | Change from baseline [95% CI] | Effect size [95% CI] | Adjusted P |
| --- | --- | --- | --- | --- | --- | --- |
| Exercise | Global Work Index | 1412.97 ± 287.40 | 1807.48 ± 381.54 | 394.52 (253.53, 535.50) | 1.026 (0.584, 1.458) | **<0.001\*\*** |
| | Peak Systolic Strain | 14.93 ± 2.01 | 17.27 ± 3.12 | 2.34 (1.37, 3.30) | 0.884 (0.462, 1.295) | **<0.001\*\*** |
| | Strain Range | 14.79 ± 1.98 | 17.35 ± 2.52 | 2.56 (1.70, 3.42) | 1.098 (0.644, 1.540) | **<0.001\*\*** |
| | End-Systolic Pressure | 109.05 ± 13.05 | 132.39 ± 19.74 | 23.34 (16.53, 30.15) | 1.257 (0.767, 1.748) | **<0.001\*\*** |
| | Systolic Strain Rate | 0.72 ± 0.12 | 0.98 ± 0.21 | 0.26 (0.18, 0.33) | 1.292 (0.807, 1.766) | **<0.001\*\*** |
| | Arterial Elastance | 7.49 ± 1.30 | 7.80 ± 1.66 | 0.31 (-0.24, 0.87) | 0.207 (-0.151, 0.561) | 0.259 |
| Handgrip | Global Work Index | 1683.77 ± 275.87 | 1763.17 ± 265.92 | 79.40 (-3.93, 162.73) | 0.356 (-0.016, 0.722) | 0.183 |
| | Peak Systolic Strain | 18.75 ± 2.67 | 18.19 ± 2.22 | -0.57 (-1.23, 0.09) | -0.322 (-0.687, 0.047) | 0.183 |
| | Strain Range | 18.35 ± 2.53 | 17.55 ± 2.27 | -0.80 (-1.57, -0.03) | -0.390 (-0.759, -0.016) | 0.165 |
| | End-Systolic Pressure | 106.14 ± 8.32 | 115.17 ± 11.59 | 9.03 (6.00, 12.06) | 1.112 (0.649, 1.563) | **<0.001\*\*** |
| | Systolic Strain Rate | 0.82 ± 0.10 | 0.80 ± 0.12 | -0.02 (-0.06, 0.02) | -0.196 (-0.573, 0.181) | 0.183 |
| | Arterial Elastance | 5.89 ± 0.98 | 6.70 ± 1.26 | 0.81 (0.43, 1.19) | 0.791 (0.375, 1.197) | **<0.001\*\*** |
| PLR | Global Work Index | 1683.77 ± 275.87 | 1767.90 ± 213.09 | 84.13 (-11.91, 180.17) | 0.327 (-0.043, 0.692) | 0.186 |
| | Peak Systolic Strain | 18.75 ± 2.67 | 17.76 ± 2.06 | -0.99 (-1.79, -0.20) | -0.465 (-0.838, -0.084) | 0.066 |
| | Strain Range | 18.35 ± 2.53 | 17.58 ± 1.88 | -0.77 (-1.57, 0.04) | -0.355 (-0.721, 0.017) | 0.186 |
| | End-Systolic Pressure | 106.14 ± 8.32 | 114.75 ± 8.31 | 8.61 (5.95, 11.27) | 1.207 (0.728, 1.674) | **<0.001\*\*** |
| | Systolic Strain Rate | 0.82 ± 0.10 | 0.79 ± 0.08 | -0.02 (-0.06, 0.02) | -0.228 (-0.589, 0.136) | 0.222 |
| | Arterial Elastance | 5.89 ± 0.98 | 6.62 ± 0.99 | 0.73 (0.37, 1.08) | 0.766 (0.353, 1.170) | **0.001\*\*** |

Exercise significantly increased all contractility-related parameters: systolic strain rate, GWI, peak systolic strain, strain range, and end-systolic pressure (all adjusted p<0.001). Arterial elastance remained unchanged, indicating a contractility-dominant response (**Supplementary Table S4**).

In contrast, handgrip and PLR primarily increased afterload indices: both interventions significantly elevated end-systolic pressure and arterial elastance (adjusted p≤0.001), while effects on GWI, strain range, peak systolic strain, and systolic strain rate were small and not significant after multiplicity correction (**Supplementary Tables S5 and S6**).

### Sensitivity Analysis

Only three outliers were identified across all 18 endpoint–intervention combinations. The nine endpoints retaining significance after Holm-Bonferroni correction showed complete robustness across all sensitivity methods, with significance persisting after outlier removal, winsorization, and permutation testing, and bootstrap confidence intervals excluding zero in all cases.

Two endpoints lost statistical significance following correction: PLR peak systolic strain (adjusted p=0.066) and handgrip strain range (adjusted p=0.165). However, both showed consistent directional effects across all sensitivity analyses, suggesting that the lack of significance after adjustment could reflect the conservative correction rather than the absence of physiological response (**Supplementary Table S7**).

### Secondary Endpoints

Secondary endpoints were analyzed exploratorily without multiplicity correction; p-values are nominal and findings hypothesis-generating. Effect sizes guided interpretation across four domains: hemodynamics, myocardial work, loop morphology, and diastolic timing (**Figure 1; Supplementary Table S8**).

**Figure 1.**
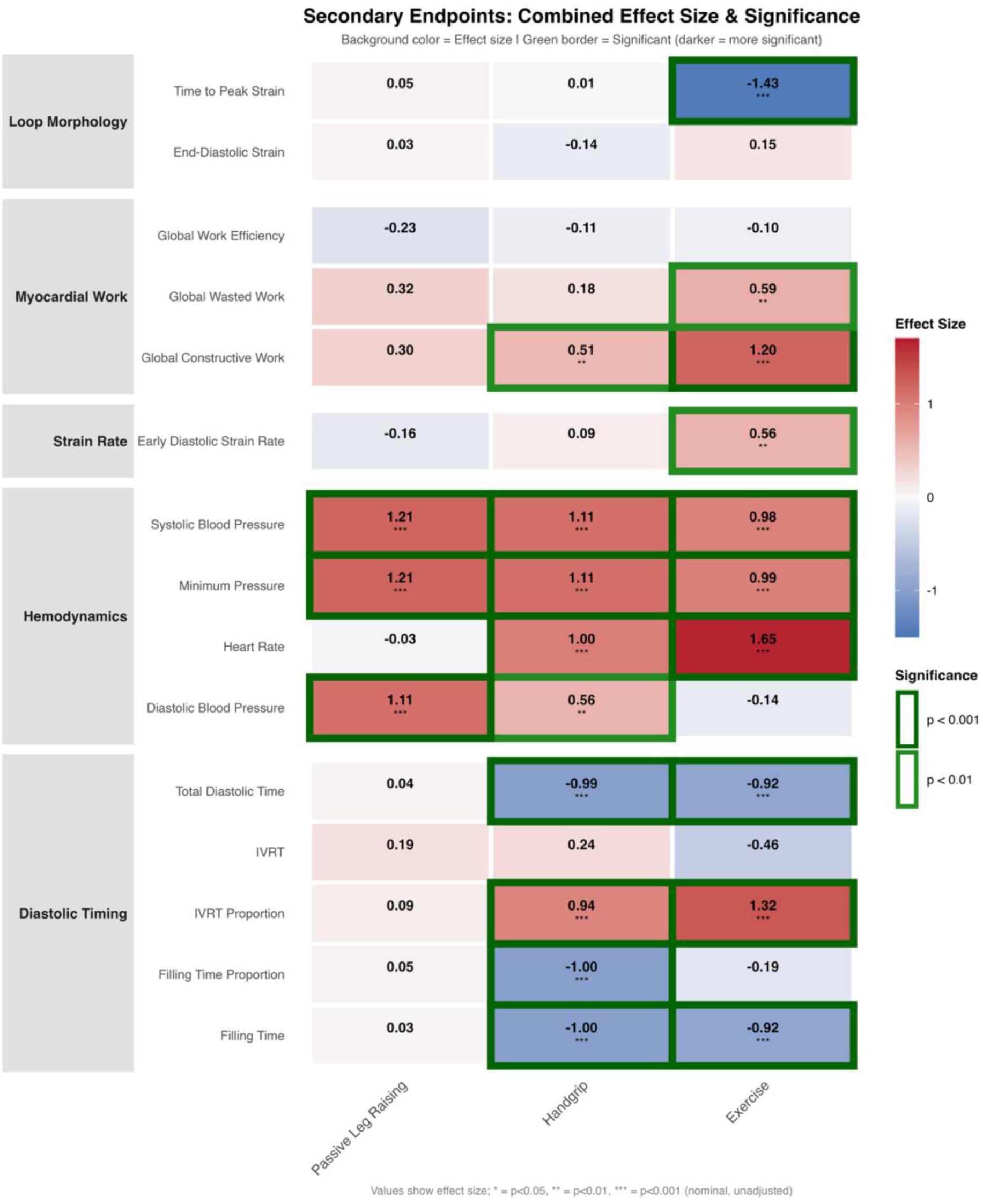
Exploratory Analysis of Secondary Endpoints: Combined Effect Size and Significance Heatmap. Heatmap visualization of exploratory secondary endpoint analyses across three hemodynamic interventions (Exercise, Handgrip, Passive Leg Raising). Cell background color represents Cohen’s d effect size magnitude (red = positive effects, blue = negative effects; color intensity proportional to |dz|). Green borders indicate nominal statistical significance: darker borders denote p<0.001, brighter borders denote p<0.01(all unadjusted for multiple comparisons). Values within cells show effect sizes with significance markers (** p<0.01, *** p<0.001, nominal). Endpoints are organized by physiological category: Loop Morphology (temporal strain characteristics), Myocardial Work (global work indices), Strain Rate (early diastolic relaxation), Hemodynamics (blood pressure and heart rate), and Diastolic Timing (IVRT and filling time proportions). **Abbreviations:** IVRT (isovolumic relaxation time)

Exercise produced the largest effects across domains, particularly for heart rate, systolic blood pressure, time to peak strain, global constructive work, and isovolumic relaxation time (IVRT) proportion, with reciprocal shortening of absolute diastolic filling times. Handgrip showed large effects primarily for heart rate and blood pressure, with diastolic timing changes secondary to rate acceleration. PLR effects were predominantly limited to blood pressure increases, with minimal impact on myocardial work or temporal parameters.

Superimposed LV-PSL illustrates changes in ventriculo-arterial coupling across conditions, with morphological shifts consistent with those reported from invasive pressure–volume catheterization (**Figure 2**).

**Figure 2.**
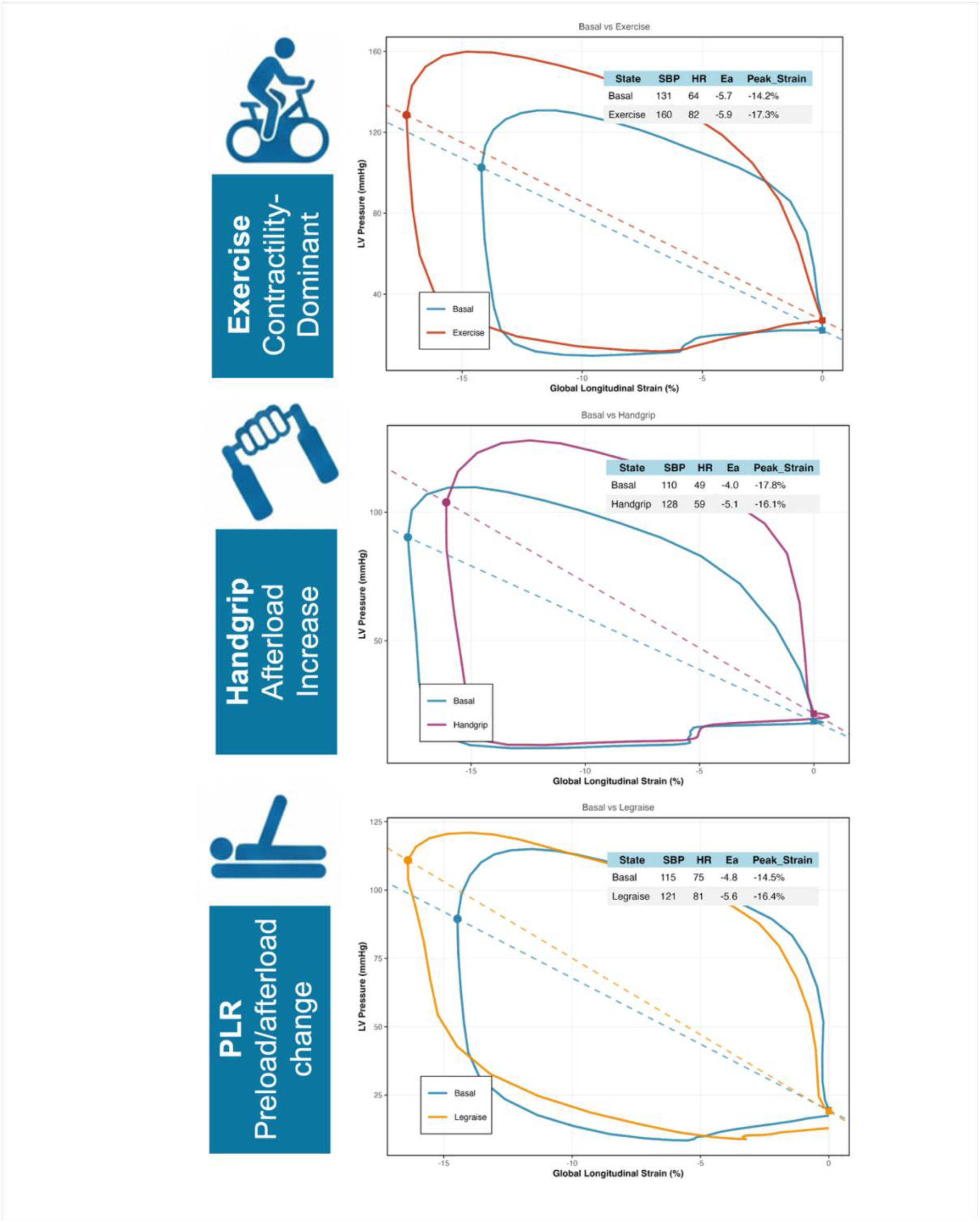
Pressure–Strain Loop Responses: Examples of Three Hemodynamic Interventions in a Healthy Volunteer. Representative pressure-strain loops at baseline (blue) and during intervention (red/purple/orange) for three distinct hemodynamic challenges. **Top panel:** Exercise (contractility-dominant response) demonstrates marked rightward and upward shift of the loop with increased area, reflecting enhanced myocardial work. **Middle panel:** Isometric handgrip (afterload increase) shows upward shift with minimal horizontal displacement, indicating preserved contractility against increased vascular resistance. **Bottom panel:** Passive leg raising (preload/afterload change) produces intermediate loop modifications with both horizontal and vertical components. Dashed gray lines represent arterial Elastance (Ea) = ESP/ Δε. Numerical values show systolic blood pressure (SBP), heart rate (HR), Ea (arterial elastance) and peak strain at baseline and during each intervention. ***Abbreviations:*** Δε, strain range; ESP, end systolic pressure; LV, left ventricular; PLR, passive leg raising

### Intra- and interobserver reliability

Fifteen participants were randomly selected from the main cohort for intra- and interobserver reproducibility analysis. Intraclass correlation coefficients (ICC) demonstrated good to excellent intra-observer reliability across all strain-derived parameters. Coefficients of variation ranged from 7.1% for peak systolic strain to 10.6% for global work index. Inter-observer reliability showed good agreement for all except for GWI, which was only moderate (see **Figure 3**). Bland–Altman plots showed small mean differences without statistically significant bias (p ≥0.25, for mean bias) and no evidence of proportional bias (p ≥0.18, for proportional bias). Plots, together with the full numerical reproducibility metrics, are presented in the **Supplementary Figures S1 and S2.**

**Figure 3.**
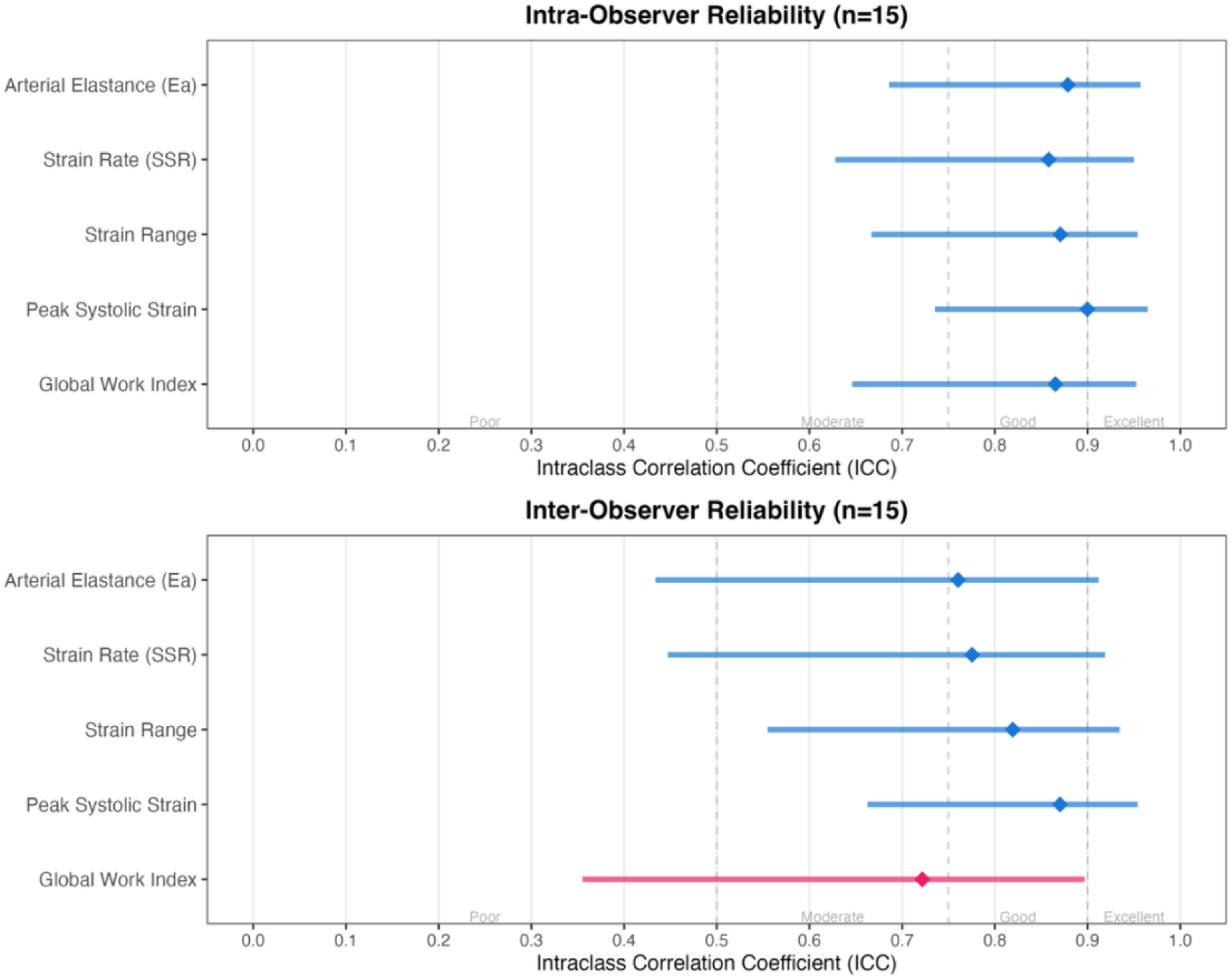
Intra- and Interobserver Reliability of Pressure-Strain Loop Parameters Intraclass correlation coefficients (ICC) with 95% confidence intervals for strain-derived parameters in 15 participants. ICC values are color-coded: poor (<0.5), moderate (0.5-0.75), good (0.75-0.90), excellent (>0.90). *Abbreviations:* SSR (systolic strain rate)

Responses across the six primary endpoints reported in the **Supplementary Figure S3 and Supplementary Table S11**, were heterogeneous. Systolic strain rate showed a consistent large increase (r=0.91, 82% responders), whereas GWI paradoxically decreased (r=−0.88). Changes in arterial elastance, end-systolic pressure, strain range, and peak systolic strain were small or negligible, with confidence intervals spanning zero.

## Discussion

In this proof-of-concept study, we evaluated non-invasive LV-PSL as a pragmatic surrogate for PV loops assessment of ventricular–arterial coupling during controlled hemodynamic stressors. Using only standard apical 2D echocardiographic views and brachial cuff pressure, LV-PSL-derived indices were able to track within-subject responses to isometric handgrip, PLR and semi-supine exercise. Across primary interventions, the direction and magnitude of changes in afterload, deformation, and myocardial work were broadly consistent with *a priori* hypotheses derived from invasive PV loops literature (15–17,20), supporting the physiological validity of LV-PSL-based metrics as non-invasive surrogates of VA coupling dynamics.

In contrast to conductance-catheter PV analysis, which remains the reference standard but is invasive and resource-intensive (7,21), the present protocol demonstrates that LV-PSL-based assessments can be implemented in a routine echocardiography setting.

The most coherent and robust pattern emerged during exercise. Five of six co-primary endpoints (systolic strain rate, peak systolic strain, strain range, end-systolic pressure, and GWI) exhibited large effect sizes (Cohen’s dz 0.9–1.3) and remained significant after Holm–Bonferroni correction. Arterial elastance showed no significant change. The pattern of marked augmentation of contractility-sensitive strain indices and myocardial work without a corresponding rise in afterload, aligns with PV-based descriptions of dynamic exercise, in which enhanced inotropy and heart rate increase end-systolic ventricular elastance and stroke work while effective arterial load remains relatively stable (17,22).

Secondary analyses further refine this interpretation. Exercise produced large effect-size increases in heart rate, systolic blood pressure, and global constructive work, alongside a medium-to-large effect-size rise in early diastolic strain rate and marked shortening of time to peak strain. IVRT proportion showed a medium effect-size increase, while absolute filling time and total diastolic time decreased markedly; filling-time proportion and end-diastolic strain showed negligible changes. Collectively, these findings indicate a contractility-dominant response with preserved or enhanced relaxation kinetics despite diastolic time compression, consistent with the expected prioritization of pump performance during dynamic exercise (22).

Isometric handgrip was selected as a predominantly afterload-modulating maneuver, and PLR as a preload challenge, based on established physiological data (6,7). In practice, both interventions produced large increases in end-systolic pressure and Ea (Cohen’s dz 0.8–1.2), while strain and work-based co-primary endpoints showed only small effect sizes and did not survive correction for multiple comparisons. For handgrip, this afterload-dominant profile was expected; for PLR, it reflects methodological considerations discussed below.

For handgrip, secondary endpoints confirmed a pressure-driven response: systolic blood pressure and heart rate increased markedly (both large effect sizes), diastolic pressure rose to a lesser extent, and global constructive work showed a moderate increase without a change in global work efficiency. Loop morphology and diastolic timing changes were small and explained by the heart rate acceleration. These findings are in agreement with previous reports that isometric handgrip acutely raises arterial load and blood pressure with only limited modification of intrinsic contractility in healthy individuals (23).

PLR was intended as a reversible preload challenge. However, volunteers supported their legs on a platform with knee flexion rather than undergoing fully operator-mediated elevation. This configuration likely combined venous return augmentation with increased peripheral resistance. The data support this interpretation: systolic and diastolic blood pressure, end-systolic pressure, and Ea all increased substantially, whereas heart rate, deformation indices, and myocardial work parameters changed little. Although classical PLR studies describe an “auto-bolus” of approximately 300 mL that increases preload and stroke volume without adding net fluid or substantially raising afterload (24). In our study, concomitant afterload augmentation appears to have partially offset Frank–Starling–mediated increases in deformation and work, resulting in a more attenuated response. The absence of significant changes in strain range, peak strain, GWI, and GCW, reinforces this interpretation.

Beyond directionality, the robustness and reproducibility of pressure-strain loop metrics are critical if they are to be deployed for serial monitoring. Nine of eighteen intervention-endpoint combinations met the Holm–Bonferroni threshold. All significant findings remained stable across sensitivity analyses - including outlier removal, winsorization, permutation testing, and bootstrap confidence intervals - with 94% of intervention-endpoint pairs showing concordant results.

Measurement reliability was similarly encouraging. Intra-observer and inter-observer intraclass correlation coefficients were predominantly in the good-to-excellent range, coefficients of variation ranged from 7% to 11%, and Bland–Altman analysis revealed no systematic or proportional bias. These metrics are comparable to, or slightly better than, previously reported values for global longitudinal strain and myocardial work indices (11,12,14). Importantly, the analysis relies on 2D speckle-tracking rather than 3D full-volume acquisitions, a practical advantage, as 2D imaging is faster and generally more robust in tachycardia, irregular rhythm, and dilated ventricles, where 3D datasets are often suboptimal (20,25).

Taken together, these data demonstrate that LV-PSL analysis can detect physiologically meaningful hemodynamic changes within individuals, with effect-size patterns that mirror established PV loop physiology under exercise, afterload stress, and preload modulation. This has potential relevance for settings in which repeated invasive assessment is impractical, such as day-hospital interventions, serial monitoring in hemodynamically unstable patients where robust non-invasive tools are needed (26). At the research level, the pre-registered statistical framework with explicit multiplicity control and sensitivity analyses illustrates how LV-PSL methodology can be evaluated with appropriate rigor. Whether this approach translates to populations with hypertension, cardiomyopathies, or heart failure, where myocardial work indices have shown diagnostic and prognostic value, requires dedicated validation (27).

### Limitations

Several limitations merit consideration. First, the primary cohort consisted of relatively young, healthy volunteers; generalizability to patients with advanced cardiovascular disease remains to be established. Second, although standardized, the PLR maneuver involved self-supported leg elevation with knee flexion, likely inducing mixed preload and afterload effects that precluded isolation of preload-specific responses. Third, pressure-strain loop indices were derived using a single software platform and an estimated LV pressure waveform; inter-vendor agreement and the impact of alternative pressure models remain unassessed. Finally, direct invasive pressure-volume measurements were not obtained, so validation relied on concordance with physiological principles and prior literature rather than simultaneous invasive comparison.

## Conclusion

In summary, this study provides proof-of-concept validation that non-invasively derived pressure-strain loops can track predictable shifts in ventriculo-arterial coupling induced by standardized physiological interventions. The method captured the expected dominance of contractile reserve during exercise, demonstrated sensitivity to afterload changes during handgrip, and showed good-to-excellent intra- and inter-observer reliability. These findings establish a physiological foundation for translating pressure-strain loop analysis into clinical practice, with potential applications in serial hemodynamic monitoring and individualized therapeutic guidance.

## Data Availability

The project is publicly registered on the Open Science Framework (OSF): https://doi.org/10.17605/OSF.IO/3QJGW.

## Acknowledgments

Sara Machado, MD; João Patricio, MD; Carla Costa, MD, Patricia Patrício, MD

