## Supplementary Materials for "Validation of Pressure-Strain Loops for Non-Invasive Assessment of Ventriculo-Arterial Coupling"

### Index of Supplementary Materials

**Supplementary Figure S1 -** Intra-Observer Bland-Altman Plots for Pressure-Strain Loop Parameters

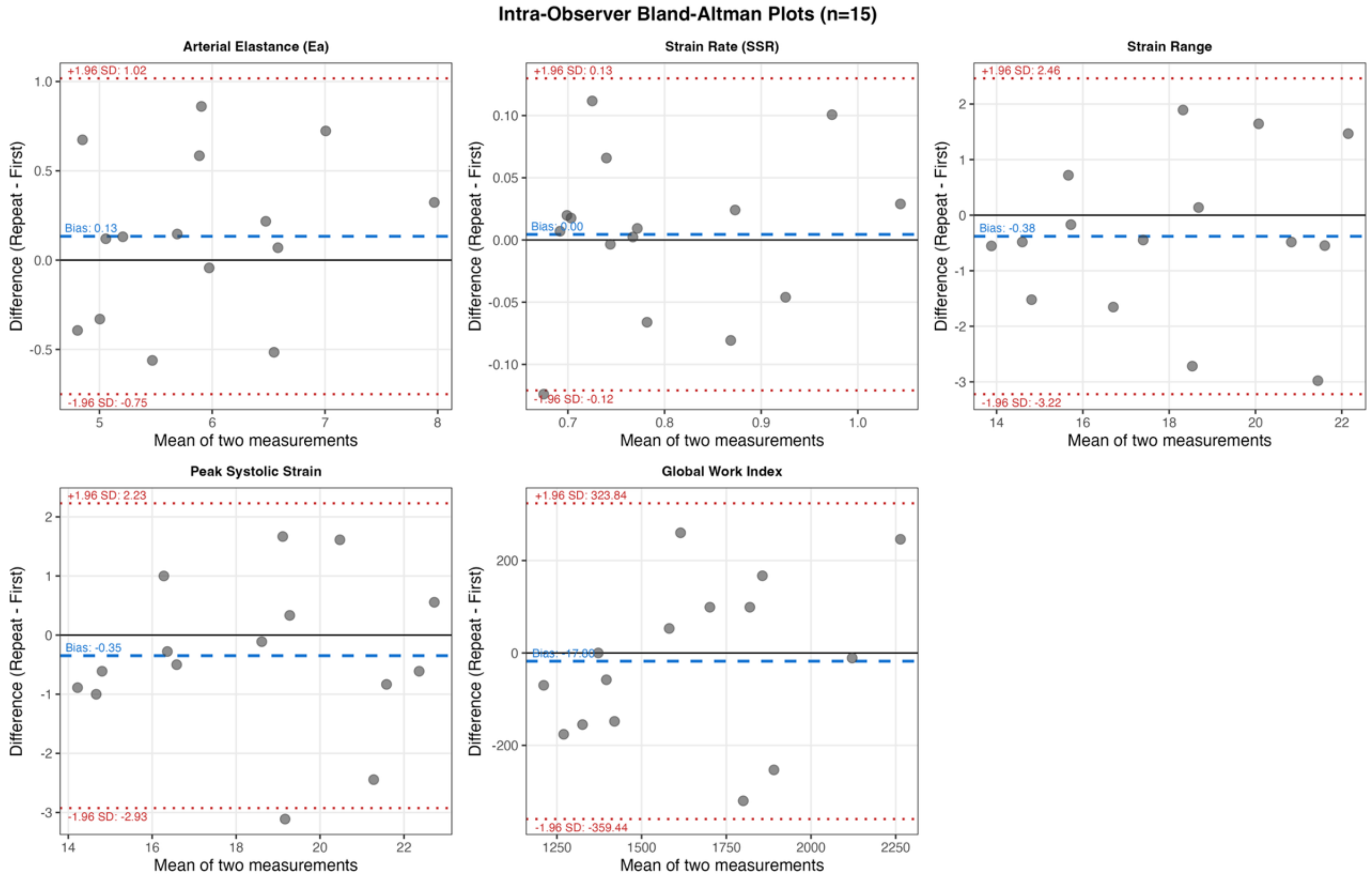

Bland-Altman plots showing intra-observer agreement for strain-derived parameters in 15 participants. The primary observer repeated offline pressure-strain loop analysis after 8 weeks. Each plot shows individual paired measurements (gray circles), mean bias (blue dashed line), and 95% limits of agreement (red dotted lines, mean ± 1.96 SD). End-systolic pressure was excluded as it did not vary between measurements. Minimal systematic bias was observed across all parameters, with acceptable measurement variability. Ea = arterial elastance; SSR = systolic strain rate; GWI = global work index.

**Supplementary Figure S2 -** Inter-Observer Bland-Altman Plots for Pressure-Strain Loop Parameters

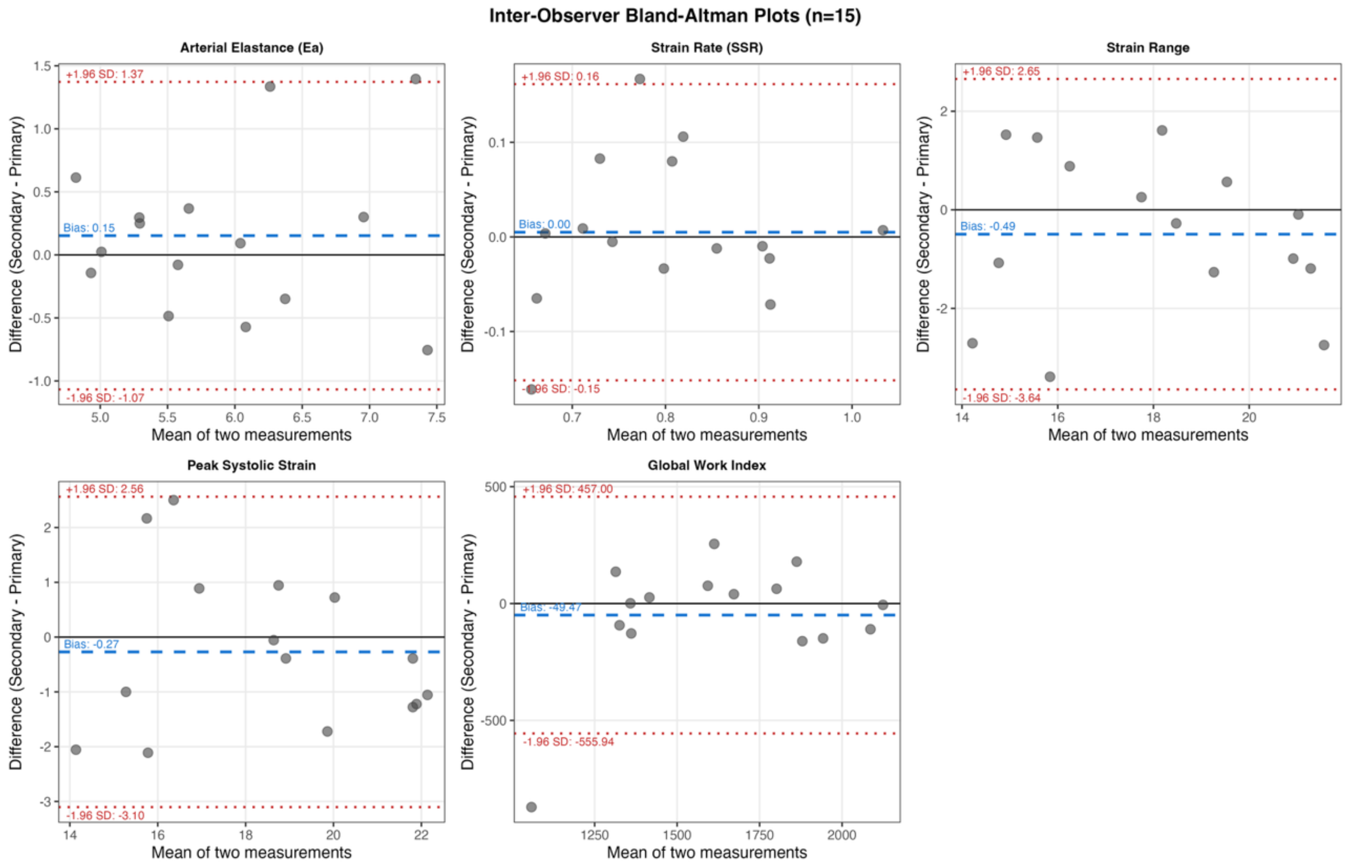

Bland-Altman plots showing inter-observer agreement for strain-derived parameters in 15 participants. Two independent observers performed offline pressure-strain loop analysis on the same echocardiographic images. Each plot shows individual paired measurements (gray circles), mean bias (blue dashed line), and 95% limits of agreement (red dotted lines, mean ± 1.96 SD). End-systolic pressure was excluded as it did not vary between measurements. Minimal systematic bias was observed across all parameters, with acceptable inter-observer measurement variability. Ea = arterial elastance; SSR = systolic strain rate; GWI = global work index

**Supplementary Table S1 -** Directional Hypotheses Based on PV Loop Physiology(1–4)

| Primary Endpoint | Dataset column | Rationale | Expected - Handgrip | Expected - PLR | Expected - Exercise |
|---|---|---|---|---|---|
| **Arterial Elastance (Ea)** | ESP/SRange_slope | Afterload | ↑ | ↑ | ↓/↔ |
| **Strain rate** | SSR_global | Contractility | ↔/↓ | ↔/↑ | ↑ |
| **End-systolic pressure** | ES_P_est | Afterload, contractility and preload | ↑ | ↑ | ↑ |
| **Loop width** | Strain_Range | Contractility, preload | ↔/↓ | ↔/↑ | ↑ |
| **Peak strain** | Peak_Systolic_strain | Contractility | ↓/↔ | ↔/↑ | ↑ (more negative) |
| **Loop area** | GWI | Contractility, afterload, preload | ↔/↑ | ↑ | ↑ |

**Note**: Arrows indicate direction of change. For strain parameters, ↓ indicates less negative values (reduced deformation), ↑ indicates more negative values (increased deformation). PLR (passive leg raising). Due to operational constraints (absence of a second operator), volunteers maintained leg elevation on a platform with knee flexion in a table-top position.

1. Patterson T, Rivolo S, Burkhoff D, Schreuder J, Briceno N, Williams R, et al. Impact of coronary artery disease on contractile function and ventricular-arterial coupling during exercise: Simultaneous assessment of left-ventricular pressure–volume and coronary pressure and flow during cardiac catheterization. Physiol Rep [Internet]. 2021 May [cited 2025 Oct 31];9(10). Available from: https://onlinelibrary.wiley.com/doi/10.14814/phy2.14768
2. Penicka M, Bartunek J, Trakalova H, Hrabakova H, Maruskova M, Karasek J, et al. Heart Failure With Preserved Ejection Fraction in Outpatients With Unexplained Dyspnea. J Am Coll Cardiol. 2010 Apr;55(16):1701–10.
3. Rommel KP, Von Roeder M, Latuscynski K, Oberueck C, Blazek S, Fengler K, et al. Extracellular Volume Fraction for Characterization of Patients With Heart Failure and Preserved Ejection Fraction. J Am Coll Cardiol. 2016 Apr;67(15):1815–25.

**Supplementary Table S2 -** Baseline Characteristics by Test in Per-Protocol Population

| Characteristic | Exercise — Total<br>N = 56[1] | Exercise — Included<br>N = 31[1] | P value | Maneuvers — Total<br>N = 47[1] | Maneuvers — Included<br>N = 30[1] | P value |
|---|---|---|---|---|---|---|
| **Sex** | | | | | | |
| Female | 32 (57%) | 20 (65%) | 0.332 | 27 (57%) | 14 (47%) | 0.093 |
| Male | 24 (43%) | 11 (35%) | 0.161 | 20 (43%) | 16 (53%) | 0.292 |
| **Age (years)** | 44 ± 9 | 42 ± 8 | 0.077 | 42 ± 8 | 43 ± 8 | 0.199 |
| **Systolic BP (mmHg)** | 119 ± 15 | 122 ± 15 | 0.197 | 117 ± 18 | 121 ± 20 | 0.057 |
| **Diastolic BP (mmHg)** | 72 ± 7 | 72 ± 8 | 0.899 | 68 ± 9 | 69 ± 9 | 0.169 |
| **BMI (kg/m²)** | 28.4 ± 4.4 | 29.0 ± 4.7 | 0.264 | 25.3 ± 3.3 | 25.1 ± 3.2 | 0.479 |
| **Any medication** | 16 (29%) | 8 (26%) | 0.832 | 12 (26%) | 8 (27%) | 1.000 |

[1]n (%); Mean ± SD

***Note:***

This table presents baseline demographic and clinical characteristics stratified by intervention type (handgrip, passive leg raising, and exercise) for participants included in the per-protocol analysis.

***Abbreviations:***

Any Medication (antidepressants, Beta 2 mimetics for asthma); BMI, body mass index; BP, blood pressure; PP, per-protocol.

**Supplementary Table S3 -** Effect Sizes by Endpoint

| Endpoint | Exercise | Handgrip | PLR | Magnitude |
|---|---|---|---|---|
| **Arterial Elastance** | 0.207 | 0.791** | 0.766** | Medium |
| **End-Systolic Pressure** | 0.977** | 1.112** | 1.207** | Large |
| **Global Work Index (GWI)** | 1.026** | 0.356 | 0.327 | Large |
| **Peak Systolic Strain** | 0.884** | -0.322 | -0.465 | Large |
| **Systolic Strain Rate** | 1.292** | -0.320 | -0.228 | Large |
| **Strain Range** | 1.098** | -0.390 | -0.355 | Large |

Values are standardized effect sizes (Cohen's d) for hemodynamic manoeuvres. ** indicates $p < 0.001$ (Holm-Bonferroni corrected).
**Abbreviations:** PLR = Passive Leg Raising.

**Supplementary Table S4** - Exercise Intervention - Echocardiographic Responses

| Endpoint | Baseline (mean ± SD) | Intervention (mean ± SD) | Mean change (95% CI) | Effect size (95% CI) | Effect_Label | Effect_ Magnitude | p_value_adjusted |
|---|---|---|---|---|---|---|---|
| **Systolic Strain Rate** | 0.72 ± 0.12 | 0.98 ± 0.21 | 0.26 [0.18, 0.33] | 1.292 [0.807, 1.766] | Cohen's dz | Large | <0.001 |
| **End Systolic Pressure** | 109.05 ± 13.05 | 132.39 ± 19.74 | 23.34 [16.53, 30.15] | 1.257 [0.767, 1.748] | Rank-biserial r | Large | <0.001 |
| **Strain Range** | 14.79 ± 1.98 | 17.35 ± 2.52 | 2.56 [1.70, 3.42] | 1.098 [0.644, 1.540] | Cohen's dz | Large | <0.001 |
| **GWI** | 1412.97 ± 287.40 | 1807.48 ± 381.54 | 394.52 [253.53, 535.50] | 1.026 [0.584, 1.458] | Cohen's dz | Large | <0.001 |
| **Peak Systolic Strain** | 14.93 ± 2.01 | 17.27 ± 3.12 | 2.34 [1.37, 3.30] | 0.884 [0.462, 1.295] | Cohen's dz | Large | <0.001 |
| **Arterial Elastance** | 7.49 ± 1.30 | 7.80 ± 1.66 | 0.31 [-0.24, 0.87] | 0.207 [-0.151, 0.561] | Cohen's dz | Small | 0.259 |

Primary in response to exercise intervention (n = 31). Values are presented as mean ± standard deviation or mean change [95% confidence interval].
Effect Size: Cohen's dz for paired t-test; rank-biserial r for Wilcoxon signed-rank test.
p_value_adjusted: Holm-Bonferroni corrected p-values for multiple comparisons.
**Abbreviations:**
CI = confidence interval; GWI = global work index; SD standard deviation

**Supplementary Table S5 -** Handgrip Intervention - Echocardiographic Responses

| Endpoint | Baseline (mean ± SD) | Intervention (mean ± SD) | Mean change (95% CI) | Effect size (95% CI) | Effect_Label | Effect_ Magnitude | p_value_adjusted |
|---|---|---|---|---|---|---|---|
| **End Systolic Pressure** | 106.14 ± 8.32 | 115.17 ± 11.59 | 9.03 [6.00, 12.06] | 1.112 [0.649, 1.563] | Cohen's dz | Large | <0.001 |
| **Arterial Elastance** | 5.89 ± 0.98 | 6.70 ± 1.26 | 0.81 [0.43, 1.19] | 0.791 [0.375, 1.197] | Cohen's dz | Medium | <0.001 |
| **Strain Range** | 18.35 ± 2.53 | 17.55 ± 2.27 | -0.80 [-1.57, -0.03] | -0.390 [-0.759, -0.016] | Cohen's dz | Small | 0.165 |
| **GWI** | 1683.77 ± 275.87 | 1763.17 ± 265.92 | 79.40 [-3.93, 162.73] | 0.356 [-0.016, 0.722] | Cohen's dz | Small | 0.183 |
| **Peak Systolic Strain** | 18.75 ± 2.67 | 18.19 ± 2.22 | -0.57 [-1.23, 0.09] | -0.322 [-0.687, 0.047] | Cohen's dz | Small | 0.183 |
| **Systolic Strain Rate** | 0.82 ± 0.10 | 0.80 ± 0.12 | -0.02 [-0.06, 0.02] | -0.196 [-0.573, 0.181] | Rank-biserial r | Medium | 0.183 |

Primary endpoints in response to handgrip intervention (n = 30). Values are presented as mean ± standard deviation or mean change [95% confidence interval].
Effect Size: Cohen's dz for paired t-test; rank-biserial r for Wilcoxon signed-rank test.
p_value_adjusted: Holm-Bonferroni corrected p-values for multiple comparisons.
**Abbreviations:**
CI = confidence interval; GWI = global work index, SD standard deviation

**Supplementary Table S6 -** PLR Intervention - Echocardiographic Responses

| Endpoint | Baseline (mean ± SD) | Intervention (mean ± SD) | Mean change (95% CI) | Effect size (95% CI) | Effect_Label | Effect_ Magnitude | p_value_adjusted |
|---|---|---|---|---|---|---|---|
| **End Systolic Pressure** | 106.14 ± 8.32 | 114.75 ± 8.31 | 8.61 [5.95, 11.27] | 1.207 [0.728, 1.674] | Cohen's dz | Large | <0.001 |
| **Arterial Elastance** | 5.89 ± 0.98 | 6.62 ± 0.99 | 0.73 [0.37, 1.08] | 0.766 [0.353, 1.170] | Cohen's dz | Medium | 0.001 |
| **Peak Systolic Strain** | 18.75 ± 2.67 | 17.76 ± 2.06 | -0.99 [-1.79, -0.20] | -0.465 [-0.838, -0.084] | Cohen's dz | Small | 0.066 |
| **Strain Range** | 18.35 ± 2.53 | 17.58 ± 1.88 | -0.77 [-1.57, 0.04] | -0.355 [-0.721, 0.017] | Cohen's dz | Small | 0.186 |
| **GWI** | 1683.77 ± 275.87 | 1767.90 ± 213.09 | 84.13 [-11.91, 180.17] | 0.327 [-0.043, 0.692] | Cohen's dz | Small | 0.186 |
| **Systolic Strain Rate** | 0.82 ± 0.10 | 0.79 ± 0.08 | -0.02 [-0.06, 0.02] | -0.228 [-0.589, 0.136] | Cohen's dz | Small | 0.222 |

Primary endpoints in response to passive leg raising (PLR) intervention (n = 30). Values are presented as mean ± standard deviation or mean change [95% confidence interval].
Effect Size: Cohen's dz for paired t-test; rank-biserial r for Wilcoxon signed-rank test.
p_value_adjusted: Holm-Bonferroni corrected p-values for multiple comparisons.
**Abbreviations:**
CI = confidence interval; PLR = passive leg raising; PLR= passive leg raising; SD standard deviation

**Supplementary Table S7 -** Comprehensive Sensitivity Analysis Results

| Intervention | Endpoint | Outliers | Original p | Effect Size | Winsorized p | Permutation p | Bootstrap CI≠0 | Concordance | Verdict |
|---|---|---|---|---|---|---|---|---|---|
| HANDGRIP N=30 | Arterial Elastance | 0 | **<0.001** | 0.791 | **<0.001** | **<0.001** | Yes | 4/4 | ROBUST |
| | Systolic strain rate | 0 | 0.129 | -0.320 | 0.133 | 0.2890 | No | 0/4 | ROBUST |
| | End-syst Pressure | 0 | **<0.001** | 1.112 | **<0.001** | **<0.001** | Yes | 4/4 | ROBUST |
| | Strain Range | 0 | **0.041** | -0.390 | **0.031** | **0.0410** | Yes | 4/4 | ROBUST |
| | Peak Systolic strain | 0 | 0.088 | -0.322 | 0.069 | 0.0720 | No | 0/4 | ROBUST |
| | GWI | 1 | 0.061 | 0.356 | **0.021** | 0.0600 | No | 1/4 | **SENSITIVE** |
| PLR N=30 | Arterial Elastance | 0 | **<0.001** | 0.766 | **<0.001** | **0.0010** | Yes | 4/4 | ROBUST |
| | Systolic strain rate | 0 | 0.222 | -0.228 | 0.148 | 0.2055 | No | 0/4 | ROBUST |
| | End-syst Pressure | 0 | **<0.001** | 1.207 | **<0.001** | **<0.001** | Yes | 4/4 | ROBUST |
| | Strain Range | 0 | 0.062 | -0.355 | 0.051 | 0.0640 | No | 0/4 | ROBUST |
| | Peak Systolic strain | 0 | **0.016** | -0.465 | **0.012** | **0.0165** | Yes | 4/4 | ROBUST |
| | GWI | 0 | 0.084 | 0.327 | 0.104 | 0.0900 | No | 0/4 | ROBUST |
| EXERCISE N=31 | Arterial Elastance | 0 | 0.259 | 0.207 | 0.229 | 0.2440 | No | 0/4 | ROBUST |
| | Systolic strain rate | 0 | **<0.001** | 1.292 | **<0.001** | **<0.001** | Yes | 4/4 | ROBUST |
| | End-syst Pressure | 1 | **<0.001** | 0.977 | **<0.001** | **<0.001** | Yes | 4/4 | ROBUST |
| | Strain Range | 0 | **<0.001** | 1.098 | **<0.001** | **<0.001** | Yes | 4/4 | ROBUST |
| | Peak Systolic strain | 0 | **<0.001** | 0.884 | **<0.001** | **0.0010** | Yes | 4/4 | ROBUST |
| | GWI | 1 | **<0.001** | 1.026 | **<0.001** | **<0.001** | Yes | 4/4 | ROBUST |

Concordance represents the proportion of methods showing statistical significance (out of 4: original test, winsorization, permutation, bootstrap CI). ROBUST = all methods agree; SENSITIVE = result depends on outlier treatment. Bold p-values indicate $p<0.05$. Effect sizes are Cohen's dz for normally distributed data and rank-biserial correlation for non-normally distributed data. Bootstrap CI≠0 indicates the 95% bootstrap confidence interval excludes zero. Permutation testing used 1,999 permutations; bootstrap used 1,999 resamples.

**Abbreviations:** GWI= global work index; End-Syst Pressure= End-Systolic Pressure

**Supplementary Table S8 -** Exploratory Analysis of Secondary Endpoints

*EXPLORATORY ANALYSIS: This analysis was not pre-specified for multiple comparison correction. P-values are nominal and provided for descriptive purposes. Interpretation should focus on effect sizes and physiological patterns. Findings are hypothesis-generating and require confirmation in future studies.*

| Intervention | Category | Parameter | Baseline | Change [95% CI] | % Change | Effect Size [95% CI] | Magnitude |
|---|---|---|---|---|---|---|---|
| Handgrip | Loop Morphology | ED_strain | 0.05 ± 0.01 | 0.00 (-0.00 to 0.01) | +2.2% | -0.135 (-0.498 to 0.267) | Negligible |
| | | Time to peak Strain | 376.80 ± 27.03 | -0.93 (-6.40 to 4.54) | -0.2% | 0.008 (-0.381 to 0.395) | Negligible |
| | Myocardial Work | GCW | 2014.87 ± 311.54 | 120.13 (32.04 to 208.23) | +6.0% | 0.509 (0.124 to 0.886) | Medium |
| | | GWW | 99.43 ± 43.11 | 20.17 (-9.75 to 50.09) | +20.3% | 0.178 (-0.225 to 0.530) | Negligible |
| | | GWE | 0.95 ± 0.02 | -0.00 (-0.01 to 0.01) | -0.4% | -0.110 (-0.468 to 0.250) | Negligible |
| | Strain Rate | EDSR_global | 0.68 ± 0.36 | 0.05 (-0.13 to 0.22) | +6.9% | 0.092 (-0.307 to 0.464) | Negligible |
| | Hemodynamics | HR | 64.77 ± 10.34 | 8.20 (5.19 to 11.21) | +12.7% | 1.000 (1.000 to 1.000) | Large |
| | | SBP | 117.93 ± 9.24 | 10.03 (6.66 to 13.40) | +8.5% | 1.112 (0.649 to 1.563) | Large |
| | | DBP | 71.47 ± 8.99 | 5.50 (1.80 to 9.20) | +7.7% | 0.556 (0.166 to 0.937) | Medium |
| | | Pressure_min | 8.61 ± 0.67 | 0.73 (0.49 to 0.98) | +8.5% | 1.112 (0.649 to 1.563) | Large |
| | Diastolic Timing | IVRT | 100.23 ± 8.32 | 2.00 (-2.32 to 6.32) | +2.0% | 0.236 (-0.167 to 0.572) | Small |
| | | Filling Time | 571.83 ± 149.25 | -109.71 (-148.51 to -70.90) | -19.2% | -0.995 (-0.998 to -0.990) | Large |
| | | Total diastolic time | 672.07 ± 146.66 | -107.71 (-147.09 to -68.32) | -16.0% | -0.991 (-0.996 to -0.979) | Large |
| | | IVRT proportion | 0.11 ± 0.02 | 0.02 (0.01 to 0.02) | +15.1% | 0.941 (0.871 to 0.974) | Large |
| | | Filling time proportion | 0.59 ± 0.05 | -0.05 (-0.07 to -0.03) | -8.7% | -0.995 (-0.998 to -0.990) | Large |
| | Loop Morphology | ED_strain | 0.05 ± 0.01 | 0.00 (-0.00 to 0.00) | +3.1% | 0.028 (-0.364 to 0.412) | Negligible |

| Intervention | Category | Parameter | Baseline | Change [95% CI] | % Change | Effect Size [95% CI] | Magnitude |
|---|---|---|---|---|---|---|---|
| Passive Leg Raising | | Time to peak Strain | 376.80 ± 27.03 | 0.73 (-5.27 to 6.73) | +0.2% | 0.046 (-0.313 to 0.403) | Negligible |
| | Myocardial Work | GCW | 2014.87 ± 311.54 | 85.30 (-19.99 to 190.59) | +4.2% | 0.303 (-0.066 to 0.666) | Small |
| | | GWW | 99.43 ± 43.11 | 17.03 (-3.08 to 37.14) | +17.1% | 0.316 (-0.053 to 0.681) | Small |
| | | GWE | 0.95 ± 0.02 | -0.01 (-0.02 to 0.00) | -0.6% | -0.229 (-0.590 to 0.136) | Small |
| | Strain Rate | EDSR_global | 0.68 ± 0.36 | -0.06 (-0.19 to 0.08) | -8.3% | -0.157 (-0.515 to 0.205) | Negligible |
| | Hemodynamics | HR | 64.77 ± 10.34 | -0.17 (-2.30 to 1.97) | -0.3% | -0.029 (-0.387 to 0.329) | Negligible |
| | | SBP | 117.93 ± 9.24 | 9.57 (6.61 to 12.53) | +8.1% | 1.207 (0.728 to 1.674) | Large |
| | | DBP | 71.47 ± 8.99 | 9.13 (6.07 to 12.20) | +12.8% | 1.113 (0.650 to 1.565) | Large |
| | | Pressure_min | 8.61 ± 0.67 | 0.70 (0.48 to 0.91) | +8.1% | 1.207 (0.728 to 1.674) | Large |
| | Diastolic Timing | IVRT | 100.23 ± 8.32 | 1.13 (-1.10 to 3.37) | +1.1% | 0.191 (-0.213 to 0.539) | Negligible |
| | | Filling Time | 571.83 ± 149.25 | 2.24 (-27.86 to 32.35) | +0.4% | 0.028 (-0.330 to 0.386) | Negligible |
| | | Total diastolic time | 672.07 ± 146.66 | 3.38 (-27.26 to 34.01) | +0.5% | 0.041 (-0.317 to 0.399) | Negligible |
| | | IVRT proportion | 0.11 ± 0.02 | 0.00 (-0.00 to 0.00) | +0.8% | 0.090 (-0.269 to 0.448) | Negligible |
| | | Filling time proportion | 0.59 ± 0.05 | 0.00 (-0.01 to 0.02) | +0.4% | 0.050 (-0.309 to 0.407) | Negligible |
| Exercise | Loop Morphology | ED_strain | 0.04 ± 0.01 | 0.00 (-0.00 to 0.00) | +2.2% | 0.152 (-0.203 to 0.505) | Negligible |
| | | Time to peak Strain | 379.35 ± 32.62 | -62.13 (-78.01 to -46.25) | -16.4% | -1.435 (-1.933 to -0.924) | Large |
| | Myocardial Work | GCW | 1780.45 ± 345.72 | 567.94 (394.78 to 741.09) | +31.9% | 1.203 (0.733 to 1.662) | Large |
| | | GWW | 179.26 ± 107.17 | 76.74 (28.51 to 124.98) | +42.8% | 0.591 (0.269 to 0.794) | Medium |
| | Myocardial Work | GWE | 0.90 ± 0.04 | -0.00 (-0.02 to 0.01) | -0.5% | -0.101 (-0.453 to 0.253) | Negligible |

| Intervention | Category | Parameter | Baseline | Change [95% CI] | % Change | Effect Size [95% CI] | Magnitude |
|---|---|---|---|---|---|---|---|
| | Strain Rate | EDSR_global | 0.41 ± 0.20 | 0.17 (0.06 to 0.28) | +40.9% | 0.562 (0.179 to 0.938) | Medium |
| | Hemodynamics | HR | 73.13 ± 11.64 | 16.97 (13.19 to 20.74) | +23.2% | 1.649 (1.098 to 2.187) | Large |
| | | SBP | 121.16 ± 14.50 | 25.94 (18.37 to 33.50) | +21.4% | 0.977 (0.949 to 0.990) | Large |
| | | DBP | 71.42 ± 8.01 | -1.19 (-4.25 to 1.86) | -1.7% | -0.143 (-0.496 to 0.212) | Negligible |
| | | Pressure_min | 8.84 ± 1.06 | 1.89 (1.34 to 2.45) | +21.4% | 0.986 (0.969 to 0.994) | Large |
| | Diastolic Timing | IVRT | 101.23 ± 4.49 | -3.26 (-6.65 to 0.13) | -3.2% | -0.458 (-0.715 to -0.091) | Small |
| | | Filling Time | 463.60 ± 141.09 | -102.00 (-141.13 to -62.88) | -22.0% | -0.923 (-0.965 to -0.835) | Large |
| | | Total diastolic time | 564.83 ± 141.65 | -105.26 (-145.13 to -65.39) | -18.6% | -0.923 (-0.965 to -0.835) | Large |
| | | IVRT proportion | 0.12 ± 0.02 | 0.02 (0.02 to 0.03) | +19.3% | 1.315 (0.826 to 1.792) | Large |
| | | Filling time proportion | 0.54 ± 0.06 | -0.01 (-0.03 to 0.01) | -1.8% | -0.193 (-0.546 to 0.164) | Negligible |

† P-values are NOMINAL (unadjusted for multiple comparisons) and provided for descriptive purposes only. ‡ Effect sizes: Cohen's d for normally distributed endpoints, rank-biserial correlation for non-normal endpoints. Magnitude categories: Negligible (|dz|<0.2), Small (0.2≤|dz|<0.5), Medium (0.5≤|dz|<0.8), Large (|dz|≥0.8). Baseline values shown as mean ± SD. All analyses used paired comparisons (paired t-test or Wilcoxon signed-rank test). Color coding indicates effect size magnitude for rapid visual pattern identification. Data are organized by intervention (Exercise, Handgrip, Passive Leg Raising) then by physiological category within each intervention.

These exploratory analyses were not pre-specified in the primary analysis plan and should be interpreted as hypothesis-generating observations requiring prospective validation.

***Abbreviations:*** *ED_strain, end-diastolic strain (preload marker); Time to peak Strain, time to peak strain from aortic valve closure (ms); GCW, Global Constructive Work (mmHg·%); GWW, Global Wasted Work (mmHg·%); GWE, Global Work Efficiency (%); EDSR_global, Early Diastolic Strain Rate ($s^{-1}$); HR, Heart Rate (bpm); SBP, Systolic Blood Pressure (mmHg); DBP, Diastolic Blood Pressure (mmHg); Pressure_min, Minimum Pressure (mmHg); IVRT, Isovolumetric Relaxation Time (ms); Filling Time, Ventricular Filling Duration (ms); Total diastolic time, Total Diastolic Time (ms); IVRT proportion, IVRT as Proportion of Diastole (%); Filling time proportion, Filling Time as Proportion of Diastole (%).*
